# A foundational transformer leveraging full night, multichannel sleep study data accurately classifies sleep stages

**DOI:** 10.1101/2024.08.02.24311417

**Authors:** Benjamin Fox, Joy Jiang, Sajila Wickramaratne, Patricia Kovatch, Mayte Suarez-Farinas, Neomi A Shah, Ankit Parekh, Girish N Nadkarni

## Abstract

**Study Objectives:** To investigate whether a foundational transformer model using 8-hour, multi-channel data from polysomnograms can outperform existing artificial intelligence (AI) methods for sleep stage classification.

**Methods:** We utilized the Sleep Heart Health Study (SHHS) visits 1 and 2 for training and validation and the Multi-Ethnic Study of Atherosclerosis (MESA) for testing of our model. We trained a self-supervised foundational transformer (called PFTSleep) that encodes 8-hour long sleep studies at 125 Hz with 7 signals including brain, movement, cardiac, oxygen, and respiratory channels. These encodings are used as input for training of an additional model to classify sleep stages, without adjusting the weights of the foundational transformer. We compared our results to existing AI methods that did not utilize 8-hour data or the full set of signals but did report evaluation metrics for the SHHS dataset.

**Results:** We trained and validated a model with 8,444 sleep studies with 7 signals including brain, movement, cardiac, oxygen, and respiratory channels and tested on an additional 2,055 studies. In total, we trained and tested 587,944 hours of sleep study signal data. Area under the precision recall curve (AUPRC) scores were 0.82, 0.40, 0.53, 0.75, and 0.82 and area under the receiving operating characteristics curve (AUROC) scores were 0.99, 0.95, 0.96, 0.98, and 0.99 for wake, N1, N2, N3, and REM, respectively, on the SHHS validation set. For MESA, the AUPRC scores were 0.56, 0.16, 0.40, 0.45, and 0.65 and AUROC scores were 0.94, 0.77, 0.87, 0.91, and 0.96, respectively. Our model was compared to the longest context window state-of-the-art model and showed increases in macro evaluation scores, notably sensitivity (3.7% increase) and multi-class REM (3.39% increase) and wake (0.97% increase) F1 scores.

**Conclusions:** Utilizing full night, multi-channel PSG data encodings derived from a foundational transformer improve sleep stage classification over existing methods.

## 1 Introduction

Sleep deprivation and disorders are a public health epidemic affecting about 30% of the US population, yet only 5% have been properly diagnosed [1, 2]. These conditions disrupt daily activities, mental health, and longevity, contributing to major health issues such as cardiovascular disease, cancer, diabetes, and hypertension [3, 4]. Economic and social factors, as well as climate change, are worsening sleep conditions globally [5, 6]. Thus, there is an urgent need for tools for accurate sleep staging, event detection, and long-term monitoring to improve diagnosis and sleep habits.

Polysomnography (PSG) is the standard method for diagnosing sleep disorders. Physiological signal data, including electroencephalogram (EEG), electrooculogram (EOG), electrocardiogram (ECG), and electromyogram (EMG) are collected during a sleep study, in addition to vital signs such as respiration rate and oxygen saturation [7]. These signals are manually annotated and reviewed by a clinician where they identify sleep stages [8]. The manual annotation and review process is time and resource intensive and may limit scalability [8–10].

PSG, in addition to at home sleep studies and wearable devices, all generate high frequency physiological signal data that would benefit from a harmonized approach to the interpretation of these signals during sleep [11]. State-of-the-art AI algorithms, such as the transformer architecture [12], could provide a tool for the assessment and annotation of sleep signals throughout the night. Here, we focus on building a standardized approach to encode PSG data and as proof of concept, use the encodings to predict sleep stages.

Previous work has primarily concerned using machine learning models for specific tasks with PSG data, especially the supervised classification of sleep stages with good accuracy [13–27]. Other machine learning methods have used PSG data to predict sleep related events, such as respiratory and cardiac events [28–34]. For sleep staging, these models have limitations. First, models only utilize EEG, EOG, and/or EMG. While these signal channels may be adequate for sleep stage classification, they limit the ability of the models to be transferred to other sleep tasks (via fine tuning, probing, few shot, or zero shot learning approaches). Further, these models typically are trained on time-frequency images generated from time series signal data in thirty second epochs (sleep stages are labeled every thirty seconds, per American Academy of Sleep Medicine guidelines [7], also known as a “sleep epoch”). This limits automated full night sleep staging using a particular model because for every channel, each sleep epoch of signal data must be processed to generate a time-frequency image and passed through the model, increasing inference time. Lastly, most recent state-of-the-art sleep stage classification models do not learn long term dependencies between sleep epochs throughout an entire night’s sleep. The longest context window in a state-of-the-art model is 90 minutes [17] while others typically only utilize 30 seconds of sleep data.

Advances in time series machine learning have focused recently on self-supervised, representation learning using the transformer architecture [35–38] to develop foundation models in various time series classification and forecasting tasks. Generally, these models use low frequency data, primarily < 1 Hz, limiting their fine-tuning ability to high frequency sleep datasets. Traditionally, the transformer architecture, the backbone behind large language models, has not performed well for time series tasks [39]. However, recent advances in the transformer for time series, specifically patch time series transformer (PatchTST), time frequency consistency (TFC), and MedFormer, have made the case for utilizing the transformer in time series forecasting and classification tasks [40–42]. In the sleep domain, a foundational transformer model trained with PSG data could assist with a variety of tasks in the clinic, including automating annotations and predicting health outcomes, which would save clinicians time, reduce variability in manual scoring, and help scalability by enabling treatment of more people [10]. Further, models trained on PSG data via self-supervision could be transferred to predict multiple outcomes [43, 44].

In this work, we present a novel 8-hour, multichannel patch foundational transformer (PFTSleep) trained via masked autoregression and based on the PatchTST architecture [40]. We utilize the Sleep Heart Health Study (SHHS) [45, 46] and the Multi-Ethnic Study of Atherosclerosis (MESA) [46, 47] PSG dataset for training, validation, and testing of our model. In total, our model was trained with 587,944 hours of sleep study signal data. It is state-of-the-art in sleep stage classification and the first to effectively encode an entire night’s sleep data.

## 2 Methods

This work used deidentified, retrospective PSG data collected from multicenter cohort studies and made available through the National Sleep Research Resource (NSRR) [48]. Data access was approved for use by the NSRR.

### 2.1 Datasets

We trained PFTSleep on 8,444 sleep studies acquired from the SHHS [45, 46] visit 1 PSG dataset collected from 1995-1998 (n=5,793) and SHHS visit 2 PSG dataset collected from 2001-2003 (n=2,651). The MESA [46, 47] PSG dataset collected from 2010-2012 (n=2,055) was used as an independent test set. All studies were type II, unattended at home sleep studies.

### 2.2 Data Extraction and Preprocessing

We extracted signal channels from EDF files and stored them in zarr [49] format for more performant data loading. Specifically, the C4-M1 EEG (125 Hz), left EOG (50Hz), EMG (125 Hz), ECG (visit 1: 125 Hz, visit 2: 250 Hz), SpO2 (1 Hz), thorax respiration (10 Hz), and abdomen respiration (10 Hz) were extracted from SHHS visit 1 and SHHS visit 2. Hypnograms indicating REM and NREM (wake, N1, N2, N3) sleep were labeled for each 30 seconds of sleep data following American Academy of Sleep Medicine guidelines [7]. 30 second epochs that were labeled as unknown were ignored during training. Sleep stage 4 was truncated into stage 3. Similarly, channels were extracted from the MESA study including the C4-M1 EEG (256 Hz), Left EOG (256 Hz), EMG (256 Hz), ECG (256 Hz), SpO2 (1 Hz), thorax respiration (32 Hz), and abdomen respiration (32 Hz). Again, hypnograms were extracted, and unknown 30 second epochs were ignored during testing. Sleep studies were trimmed to 8 hours or extended to 8 hours with zero padding. All channels were resampled to 125 Hz. No additional preprocessing was performed. After resampling and padding, the length of an input sample into our model was 7×3.6 million.

### 2.3 Input Data Preparation

Each 8-hour sleep study after resampling was normalized via a learned reversible instance normalization layer [50] (sample-channel wise unit variance scaling with a learnable affine parameter) and patched into 6 second patches, effectively creating a 3D tensor with 7 channels (like an image with 3 RGB channels). Each channel’s patches were then passed through an individual linear layer to embed the 6 second patches into a vector of size 512 for input into the transformer encoder architecture. The input into the transformer is a 3D tensor of shape 7×4800×512.

### 2.4 Model Architecture

The overall approach and architecture are shown in Figure 1. De novo code was developed for the purpose of this task and used concepts from the original PatchTST implementation [40]. The transformer architecture was chosen as the encoder for its ability to learn relationships across sequence data, due to its attention mechanism and parallelized framework [12]. PatchTST performs better than the original transformer for time series tasks by patching channel inputs into intervals of time, as previously described. Patches, or intervals, of time series data (vs single time points) are more meaningful representations to encode because timepoints are meaningful in relation to nearby timepoints. We patched channel data into 6 second segments to better identify specific sleep events; in comparison, previous encoders capture 30 second segments for labeling sleep stages. Thus, our architecture learns the relationship among channel patches, and its encodings can be adapted for downstream diagnostic and prediction tasks. Additionally, the encoder included a key padding mask in the attention mechanism to focus on relevant parts of the input (for example, sleep studies less than 8 hours would be padded to 8 hours, but the padded values would be ignored in attention). This allows for translatability to varying length sleep studies and sleep studies without all 7 channels that the original model was trained on. This differs from the original PatchTST architecture, which used far shorter patch and sequence lengths, a different masking strategy, and no key padding mask in attention. For sleep stage classification of each 6 second patch, a bidirectional gated recurrent unit (GRU) was used. Bidirectional GRUs do not suffer from the vanishing gradient problem like traditional recurrent neural networks and are less complex than long short-term memory networks. Model architecture and training were performed using PyTorch and Lightning. See supplementary methods and Supplementary Figure 1 for training details.

**Figure 1:**
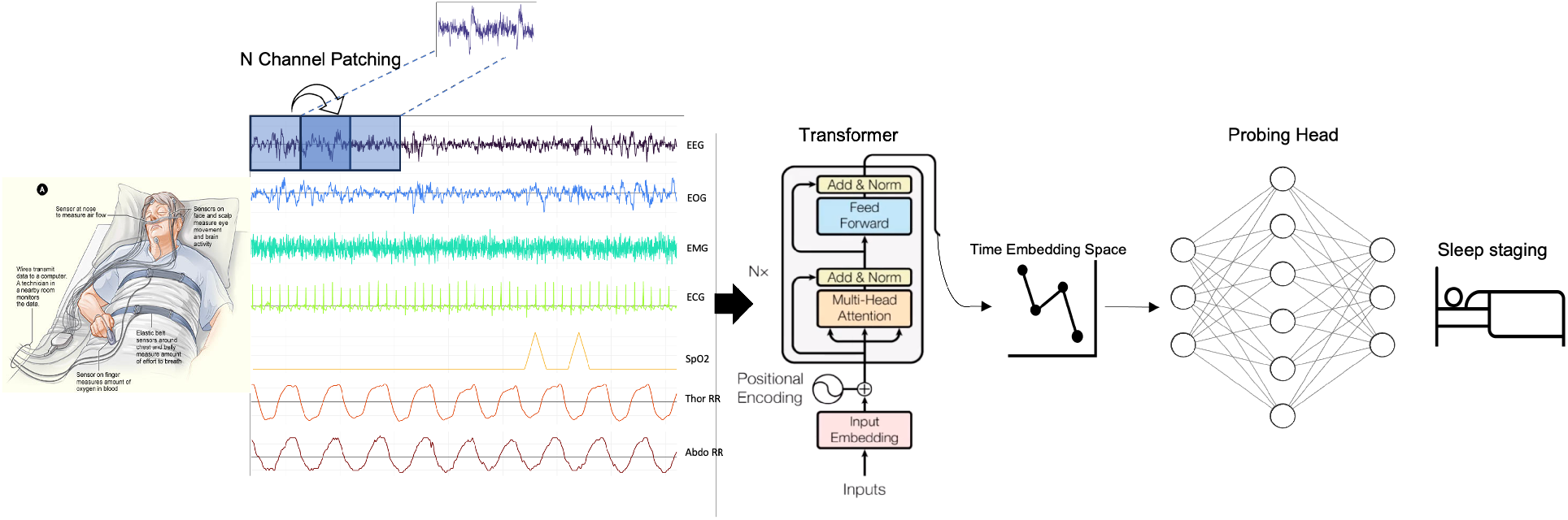
Model architecture overview. A patched (6 seconds) based self-supervised transformer model learns representations of EEG, EOG(L), EMG, ECG, SpO2, abdomen RR, and thoracic RR signals. Following, probing heads are trained to predict outcomes in sleep.

### 2.5 Statistical Analysis and Evaluation

Model performance was evaluated with macro accuracy, sensitivity, specificity, F1 scores, and Cohen’s Kappa for comparison to other models on the full training and validation, validation only, and testing only datasets. Comparisons to other models are not direct, as training and validation sets differ. Sleep stage classification is inherently an imbalanced classification problem, and metrics reported should account for this [51]. Therefore, we include area under the precision recall curves (AUPRC) to examine precision and recall over multiple decision thresholds. Additionally, area under the receiver operating characteristics curves (AUROC) and confusion matrices are reported for the validation and test sets.

## 3 Results

PFTSleep is a foundational transformer trained and tested with 587,944 hours of sleep study data. Demographics of the studies, training, and validation sets are presented in Table 1. The median age for the training, validation, and testing were 65, 66, and 68 years, respectively. The age range of the MESA dataset was 54-94 years, while it was 39-90 for SHHS. Stage 1 sleep was the lowest proportion of sleep across all data sets. Wake and stage 1 sleep in MESA were notably increased in comparison to the SHHS training and validation sets, while stage 3 had the opposite effect.

**Table 1:** Demographics and sleep stage proportions of SHHS visits 1 and 2, MESA, and SHHS training and validation sets.

|  | n | Median Age<br>(IQR) | Self-Reported<br>Gender,<br>% Female | Wake | N1 | N2 | N3 | REM |
| --- | --- | --- | --- | --- | --- | --- | --- | --- |
| SHHS Visit 1 | 5793 | 63 (55-72) | 52.4% | 27.2% | 3.7% | 41.8% | 13.3% | 14.1% |
| SHHS Visit 2 | 2651 | 68 (61-77) | 54.5% | 30.4% | 3.6% | 40.2% | 11.9% | 13.9% |
| MESA | 2055 | 68 (62-76) | 53.6% | 35.2% | 8.5% | 37.8% | 7.2% | 11.2% |
| SHHS Training Set | 6741 | 65 (57-74) | 53.2% | 28.2% | 3.7% | 41.2% | 12.9% | 13.9% |
| SHHS Validation Set | 1703 | 66 (57-74) | 53.4% | 28.2% | 3.7% | 41.5% | 12.5% | 14.1% |

The best encoder model had the lowest validation MSE loss on epoch 28. While it is hard to assess whether an encoder effectively captures signal data without testing encodings on downstream tasks, recreations of a 10 second sample of signal data are shown in Figure 2. Recreated signals are smoothed out in comparison to the originals. For sleep stage classification, numerous probing heads were explored, including multi-layer perceptron, transformer decoders, and convolutional heads. It was found that a bidirectional GRU performed the best on the sleep stage classification task.

**Figure 2:**
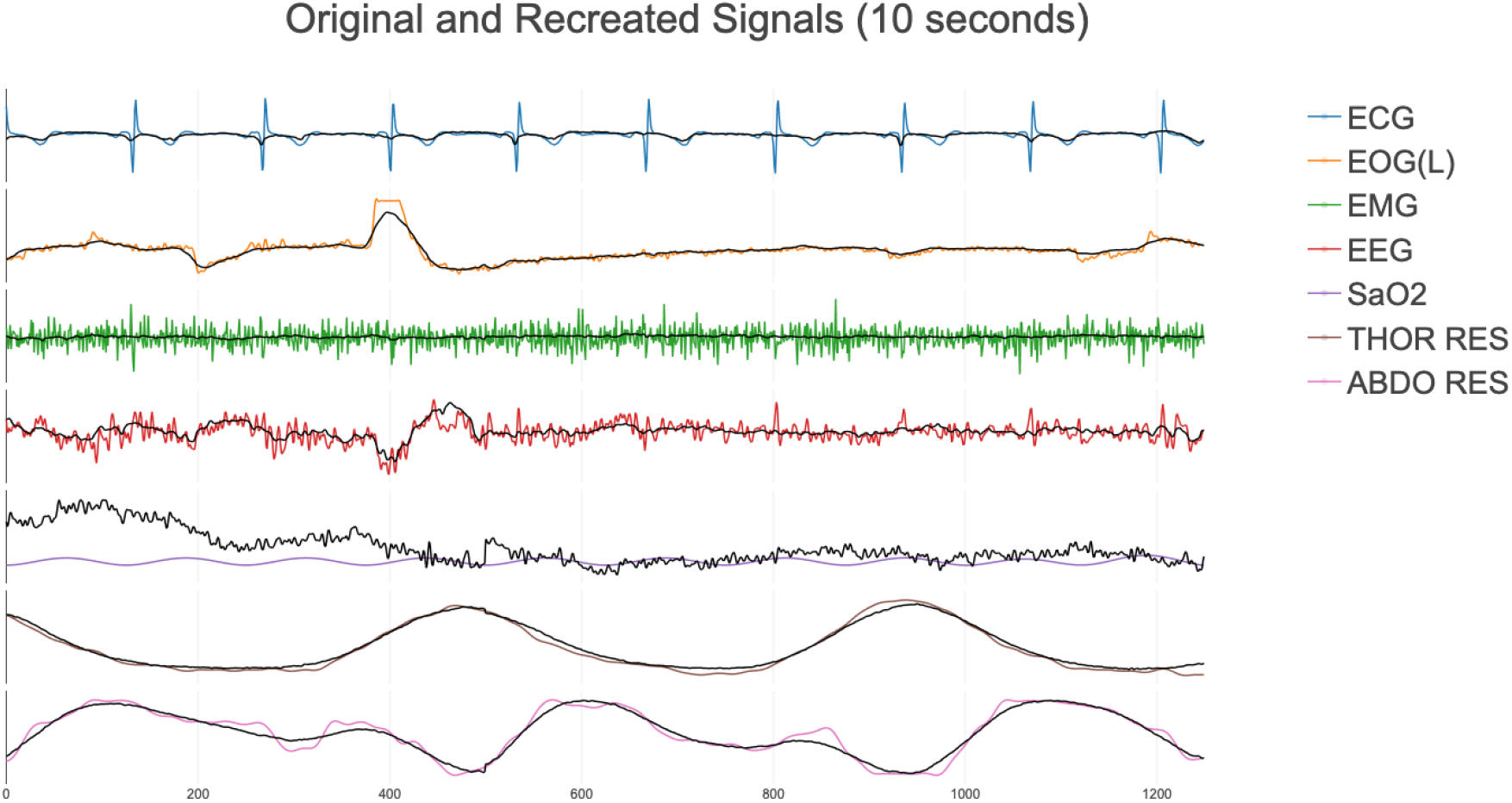
Reconstructed channels for 10 second patch of signal data after training foundational transformer model. Recreated values are in black.

### 3.1 Sleep Stage Classification Results

Overall, the AUPRC scores for wake, N1, N2, N3, and REM sleep on the SHHS validation set (the 20% not used in training) were 0.70, 0.40, 0.53, 0.75, and 0.82, respectively. Compared to the proportions of the classes in the training dataset in Table 1, the model can effectively identify instances of the positive class better than a random model. In comparison to other models, PFTSleep is state-of-the-art and performs consistently high across the 5 sleep stages for the SHHS validation set and full SHHS dataset. Table 2 shows the performance of PFTSleep in relation to other models trained on SHHS. We compared our model to the current longest context window, state-of-the-art model: L-SeqSleepNet [17]. While L-SeqSleepNet did not report AUPRC scores, our model showed marked increases in macro sensitivity (3.65% increase), and REM (3.39% increase) and wake (0.97% increase) F1 scores and was matched across other macro evaluation scores. AUROC scores were 0.99, 0.95, 0.96, 0.98, and 0.99 for wake, N1, N2, N3, REM, respectively and displayed in Table 3. Confusion matrices for the SHHS validation set are reported in Figure 3.

**Table 2:** Sleep stage evaluation metrics in comparison to state of the art models on SHHS, the full dataset, and the 20% validation set. Table adapted from Phan et al. [17]. Note that SHHS sets may not be identical, and we compared to the full SHHS dataset. Bold indicates best performance.

| Model | Overall Performance |  |  |  |  | Class Wise F1 |  |  |  |  |
| --- | --- | --- | --- | --- | --- | --- | --- | --- | --- | --- |
|  | Acc | K | MF1 | Sens | Spec | Wake | NI | N2 | N3 | REM |
| PFTSleep (SHHS full) | **87.70** | **0.83** | 80.80 | **82.30** | **96.70** | 93.30 | 48.60 | 87.80 | 82.70 | 91.50 |
| PFTSleep (SHHS val) | 85.77 | 0.80 | 0.79 | 80.25 | 96.22 | 92.50 | 45.87 | 85.90 | 78.61 | 90.09 |
| L-SeqSleepNet [17] | 87.60 | 0.83 | 80.30 | 79.40 | 96.50 | 92.40 | 48.60 | 88.20 | 83.90 | 88.50 |
| SeqSleepNet [19] | 87.20 | 0.82 | 80.20 | 78.70 | 96.30 | 91.80 | 49.10 | 88.20 | 83.50 | 88.20 |
| SleepTransformer [22] | 87.70 | 0.83 | 80.10 | 78.70 | 96.50 | 92.20 | 46.10 | 88.30 | **85.20** | 88.60 |
| XSleepNet1 [27] | 87.60 | 0.83 | 80.70 | 79.70 | 96.50 | 91.60 | **51.40** | **88.50** | 85.00 | 88.40 |
| XSleepNet2 [27] | 87.50 | 0.83 | **81.00** | 80.40 | 96.50 | 92.00 | 49.90 | 88.30 | 85.00 | 88.20 |
| U-Sleep [23] | - | - | 80.00 | - | - | 93.00 | 51.00 | 87.00 | 76.00 | **92.00** |
| Olesen et al. [25] | 87.10 | 0.82 | 78.80 | 77.70 | 96.30 | **94.10** | 47.80 | 87.90 | 74.30 | 89.90 |
| CNN [13] | 86.80 | 0.81 | 78.50 | - | 95.00 | - | - | - | - | - |
| FCNN+RNN [27] | 86.70 | 0.81 | 79.50 | 78.10 | 96.20 | 91.10 | 48.70 | 88.00 | 82.60 | 87.10 |
| IITNet [16] | 86.70 | 0.81 | 79.80 | - | - | - | - | - | - | - |
| AttnSleep [26] | 84.20 | 0.78 | 75.30 | - | - | 86.70 | 33.20 | 87.10 | 87.10 | 82.10 |

**Table 3:** AUROC and AUPRC scores for the SHHS validation set and MESA test sets.

|  | Average |  | Class wise AUROC |  |  |  |  | Class wise AUPRC |  |  |  |  |
| --- | --- | --- | --- | --- | --- | --- | --- | --- | --- | --- | --- | --- |
|  | AUROC | AUPRC | Wake | N1 | N2 | N3 | REM | Wake | N1 | N2 | N3 | REM |
| PFTSleep (SHHS val) | 0.97 | 0.66 | 0.99 | 0.95 | 0.96 | 0.98 | 0.99 | 0.82 | 0.40 | 0.53 | 0.75 | 0.82 |
| PFTSleep (MESA) | 0.89 | 0.44 | 0.94 | 0.77 | 0.87 | 0.91 | 0.96 | 0.56 | 0.16 | 0.40 | 0.45 | 0.65 |

**Figure 3:**
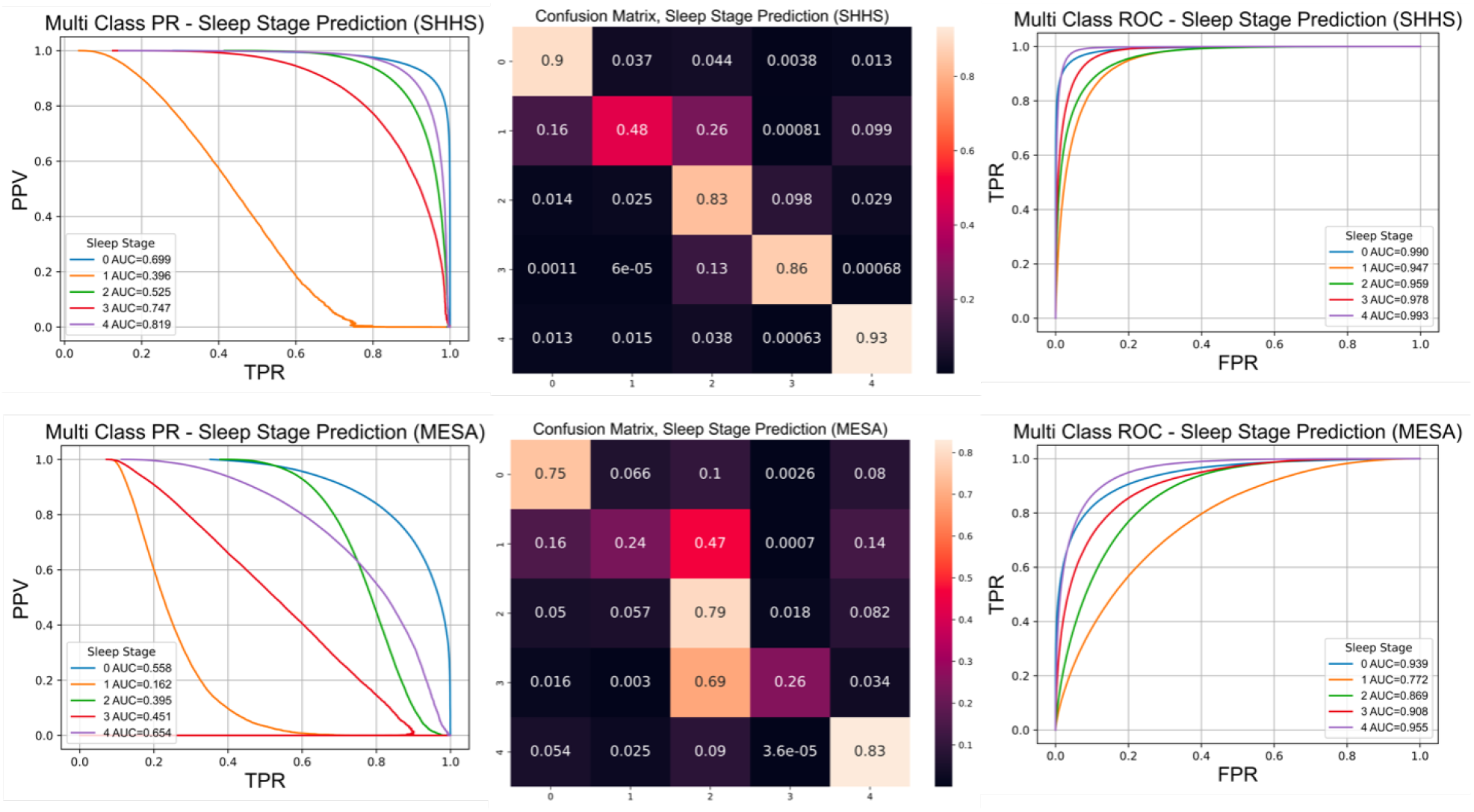
Multi class precision recall (PR) curve (first column), row normalized confusion matrix (2nd column), and multi class receiver operating characteristic (ROC) curve (third column) for sleep stage classification on 20% SHHS validation set (first row) and MESA (second row). Acronyms: FPR = false positive rate, PPV = positive predictive value, TPR = true positive rate. Confusion matrices keys: 0 = wake, 1 = N1, 2 = N2, 3 = N3, 4 = REM sleep.

### 3.2 External Testing Results

The MESA dataset was evaluated using identical metrics and reported in Table 4. Overall, the AUPRC scores for MESA were 0.56, 0.16, 0.40, 0.45, and 0.65 for wake, N1, N2, N3, REM, respectively. AUROC scores were 0.94, 0.77, 0.87, 0.91, and 0.96 and reported in Table 3 and Figure 3. Confusion matrices are illustrated in Figure 3.

**Table 4:** Sleep stage evaluation metrics on the external testing set, MESA.

|  | Overall Performance |  |  |  |  | Class Wise F1 |  |  |  |  |
| --- | --- | --- | --- | --- | --- | --- | --- | --- | --- | --- |
|  | Acc | K | MF1 | Sens | Spec | Wake | N1 | N2 | N3 | REM |
| PFTSleep | 69.57 | 0.56 | 56.97 | 57.27 | 91.50 | 80.43 | 26.19 | 73.70 | 37.51 | 67.02 |

## 4 Discussion and Conclusion

We present a novel foundational transformer, PFTSleep, that predicts sleep stages with state-of-the-art performance. It is the first PSG based model to input an entire night’s sleep - 8 hours of sleep signal data including brain, movement, cardiac, oxygen, and respiratory channels. The supervised probing head can effectively predict sleep stages for the entire night from the encodings of the foundational transformer with a single step. Performance was state-of-the-art for the SHHS validation set. While performance decreased on the MESA dataset, we hypothesize that age differences particularly in N3 (deep sleep) could be a contributing factor, and additional training data could improve performance.

To date, only one sleep stage classification model has used a transformer architecture, SleepTransformer, but the input is a time-frequency image of a 30 second segment of EEG, EOG, and/or EMG signal data, and its primary purpose is to predict sleep stages [22]. For a single 8-hour sleep study, this would require 960 inference steps, in addition to the preprocessing required to generate spectrogram images for each channel used in the model. PFTSleep can annotate a full sleep study with one inference step and minimal preprocessing (resampling, instance normalization, and patching). Another study, SleepFM [43], built a novel foundational convolutional encoder and showed good performance on sleep stage classification and sleep disordered breathing across multiple brain, cardiac, and respiratory channels. Again, the input to the model was 30 seconds worth of sleep data, potentially limiting the model’s ability to capture specific sleep dynamics throughout the night. Their encoder learned relationships via a contrastive loss function among 30 second embeddings of brain, cardiac, and respiratory channels. During training, brain, cardiac, and respiratory channels were separated into their own respective encoders and embeddings are compared pairwise and via a leave one out method. They assessed the model on an internal test set and external test set of 100 samples from the Physionet Computing in Cardiology 2018 challenge [52]. They did not validate their model on a larger, NSRR dataset for comparison. Finally, Ogg et al. built a foundational convolutional transformer using EEG signals and a sequence length of about 1 hour, demonstrating the utility of self-supervised models over fully supervised models and the encoder’s ability to translate to downstream tasks, such as sleep stage and brain age prediction. Again, this model only used EEG data and did not model the entire night’s sleep [44]. While they did utilize NSRR sleep studies, evaluation metrics were not reported for specific tasks, limiting comparison.

PFTSleep addresses several limitations in these studies, notably the ability to encode a full night of sleep that effectively learns relationships among and within channels throughout the night. Additionally, patching channels into 6 second segments could capture more specific sleep events in comparison to encoders that model 30 second segments (which is traditionally used for labeling sleep stages). Clinically, PFTSleep could assist with annotations of sleep studies, with only a single forward pass through the model (as opposed to other models, which require inputting each 30 second sleep epoch). The GRU head also can be fine-tuned to an individual laboratory’s PSG data or outcomes of interest to the laboratory beyond sleep staging. From an industry perspective, a foundational model integrated into consumer wearables like PFTSleep could better evaluate and standardize sleep health metrics across different wearables, allowing for more transparency into how the metrics were developed and their use in research.

Our work has limitations. First, the foundational transformer encoder was trained via masked autoregression, a common training technique in imaging and more recently in time series. Evaluating the performance of masked autoregressive training is difficult, without further training on multiple downstream tasks. Visualizing recreated encodings should be interpreted cautiously as they are recreated through a single linear layer. Second, we trained on datasets that were at home sleep studies and did not use clinical PSG data. The NSRR has many clinical PSG datasets that can be further used for training the foundational transformer model and the GRU head. We are actively working on expanding the dataset size and testing on specific populations to ensure the model is robust. Third, we did not train and test additional heads for downstream tasks other than sleep stage classification, but this model has the potential to enable alternative tasks in sleep. Future studies could include training heads for other sleep outcomes including cardiovascular risk prediction or daytime sleepiness, among others. Finally, the novel architecture we utilized does not allow for explainability. However, recent advancements in time series explainable AI [53] and attention-based relevance propagation for transformers [54] could provide valuable insights into the specific characteristics of PSG signal data that are related to the outcome.

In summary, we developed and validated a novel foundational transformer that utilizes full night, multichannel data to accurately classify sleep stages. We envision this approach could unlock the potential of full night sleep data for a variety of biomedical research and clinical applications.

## Data Availability

All data produced in the present study are available upon reasonable request to the authors.

https://sleepdata.org/datasets/shhs/

https://sleepdata.org/datasets/mesa/

## Acknowledgments

This work was supported in part through the computational and data resources and staff expertise provided by Scientific Computing and Data at the Icahn School of Medicine at Mount Sinai and supported by the Clinical and Translational Science Awards (CTSA) grant UL1TR004419 from the National Center for Advancing Translational Sciences. Research reported in this publication was also supported by the Office of Research Infrastructure of the National Institutes of Health under award number S10OD026880 and S10OD030463. The content is solely the responsibility of the authors and does not necessarily represent the official views of the National Institutes of Health.

