## Supplemental Methods for "A foundational transformer leveraging full night, multichannel sleep study data accurately classifies sleep stages"

### 1. FOUNDATIONAL TRANSFORMER MODEL TRAINING

The initial learning rate was set to 1e-5 and a one cycle scheduler was used with a max learning rate of 0.01. The foundational transformer encoder was trained with SHHS visits 1 and 2 with 80% training and 20% validation (without patient overlap) and a 10% augmentation mask, where 10% of values across all channels are augmented with random noise or set to zero. The encoder comprised 2 layers with 2 attention heads each, a dimension of 512, and a feedforward size of 2048. The model was trained for 100 epochs with a batch size of 4 and gradient accumulation of 4 on two NVIDIA A100 80GB Tensor Core GPUs. The training objective was to minimize the mean square error (MSE) between the original, non-augmented time series and the recreated time series (from the augmented data). An overview of the training process is detailed in Supplementary Figure 1. The signal data was recreated from the embeddings through a single linear layer to compute the MSE loss. The model with the lowest MSE loss on the validation set was saved over the 100 epochs of training and used for downstream tasks.

### 2. GATED RECURRENT UNIT PROBING HEAD TRAINING

The GRU probing model was trained using the transformer encoder with the lowest MSE validation loss. A bidirectional GRU with a hidden size of 512 was trained to predict a sleep stage for every 6 second patch. Average pooling calculated a prediction for each 30 second sleep epoch (across 5 patches of data). The model was trained for 50 epochs with an initial learning rate of 1e-5 and a focal loss function, which identifies and focuses on harder to classify examples, compared to cross entropy. The focal gamma parameter was set to 2.0 and the alpha parameter was not set.

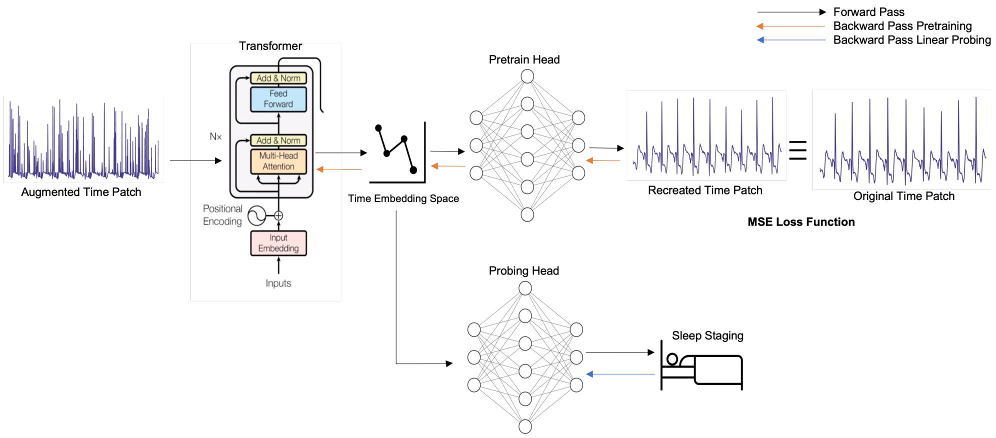


**Fig. S1.** Masked autoregression is utilized to learn the representations. Representations are used as input into probing models to predict outcomes in sleep (without adjusting weights of foundational model)
